# Carotid Artery Stenosis is Associated with Restless Legs Syndrome in the Absence of Stroke

**DOI:** 10.64898/2026.11.17.26368361

**Authors:** Jonathan Ogulnick, William Dong Jun Shin, Jason Mathew

## Abstract

**Background:** Restless legs syndrome (RLS) is a common neurological disorder with established links to atherosclerotic disease and secondary causes such as ischemic stroke. Hypothesizing that internal carotid artery stenosis (CAS) could induce secondary RLS through chronic hypoperfusion or microemboli to dopaminergic subcortical structures, even without overt stroke, we investigated the association between bilateral CAS and RLS.

**Methods:** A retrospective propensity score-matched case-control study was conducted using the TriNetX Global Health Research Network. Cases (n=58,382) had bilateral CAS/occlusion (ICD-10-CM I65.23); controls (n=58,382) had atherosclerosis without CAS. Patients were excluded for prior stroke/TIA, iron deficiency, antipsychotic use, and other secondary RLS causes. Cases and controls were 1:1 matched on age, race/ethnicity, chronic kidney disease, peripheral neuropathy, peripheral artery disease, Parkinson’s disease, and SSRI use. The outcome was RLS diagnosis (G25.81), assessed via conditional logistic regression.

**Results:** RLS was diagnosed in 1,154 (1.977%) patients with bilateral CAS/occlusion versus 1,060 (1.816%) controls, yielding an absolute risk difference of 0.161% (95% CI 0.005–0.317%; p=0.0437). The adjusted odds ratio was 1.09 (95% CI 1.002–1.186), indicating a modest but significant association.

**Conclusion:** Severe bilateral CAS/occlusion is associated with a small but statistically significant increase in RLS risk, independently of overt cerebrovascular events and known confounders. These findings support a potential vascular contribution to RLS pathogenesis, possibly through subclinical hypoperfusion affecting basal ganglia dopaminergic pathways. Prospective studies are needed to explore causality, dose-dependency, and the impact of carotid revascularization.

## INTRODUCTION

Restless legs syndrome (RLS) and carotid artery stenosis (CAS) are common neurological conditions with estimated prevalence in the United States of up to 10% and 7.4%, respectively.^1,2^ Both of these conditions can directly and indirectly cause severe quality of life impairment but are not typically discussed together in clinical and research scenarios.

The relationship between RLS and atherosclerotic disease (AD), such as peripheral artery disease (PAD), and coronary artery disease (CAD) is well established. Furthermore, it has been established that ischemic stroke can be a direct cause of secondary RLS.^3^ The shared relationship between AD and RLS is thought to be bidirectional and mediated through common pathophysiological mechanisms such as autonomic dysfunction, inflammation, and oxidative stress.^4^ One way in which RLS could contribute to AD is its relationship with both obstructive sleep apnea and periodic limb movement disorder, both of which can cause nocturnal blood pressure spikes. Both AD and RLS also share common risk factors, including obesity; smoking; poor sleep hygiene; and alcohol abuse. The bidirectional relationship is also implied by the fact that medical treatment of RLS has been linked to reduction in AD risk.^5,6^ Lastly, irony deficiency anemia has been linked to both the development of RLS at mechanistic level, and patients with AD.^7,8^

Primary RLS is thought to occur through defective dopamine synthesis in the central nervous system^1^. In the case of secondary RLS caused by stroke, Makwana et al have demonstrated that there is a strong association between development of RLS and subcortical stroke location.^9^ It is well known that dopamine is a crucial neurotransmitter in the subcortical structures of the basal ganglia.^10^ Additionally, the basal ganglia receive their blood supply primarily from branches of the middle cerebral artery (MCA) and anterior cerebral artery (ACA) - both branches of the internal carotid artery (ICA).^11^ Thus, we hypothesized that ICA stenosis even in the absence of stroke could cause secondary RLS.

Carotid stenosis contributes to subcortical infarcts not only through hemodynamic compromise or watershed mechanisms but also via artery-to-artery microembolism from unstable atherosclerotic plaques, which can occlude penetrating arteries and produce lacunar-like or multiple unilateral subcortical lesions in the ipsilateral territory. Silent subcortical brain infarcts are nearly three times more prevalent in individuals with carotid stenosis (approximately 30% vs. 17% without; pooled OR 2.78).^12^ Microembolic signals detected by transcranial Doppler occur in about 27–46% of symptomatic carotid stenosis cases (higher with near-occlusion or unstable plaques) compared to only ~6% in asymptomatic cases.^13^

To further investigate this relationship, we queried a large, global database of aggregated, deidentified electronic health records from the TriNetX network. We sought to determine whether CAS is associated with RLS in the absence of stroke or other risk factors for primary or secondary RLS in order to test our hypothesis that hypoperfusion to the basal ganglia can cause RLS. To our knowledge, this is the first study to carry out this investigation. Moreover, we have the advantage of having harnessed a large, international database.

## METHODS

### Data Source

We conducted a retrospective matched case-control study using de-identified electronic health record data from the TriNetX **Global Health Research Network** (TriNetX, LLC, Cambridge, MA, USA). This federated real-world data platform aggregates de-identified patient records from contributing healthcare organizations across multiple countries.^14^ Analyses were performed using the TriNetX online analytics platform (data accessed on December 28, 2025). No specific date range restrictions were applied for patient encounters; all available data up to the date of query were included. Note that this study used only de-identified data and was exempt from institutional review board approval.

Diagnoses, procedures, medications, and laboratory values were identified using International Classification of Diseases, 10th Revision, Clinical Modification (ICD-10-CM) codes, ICD-10-Procedure Coding System (PCS) codes, National Drug Codes (NDC), or Logical Observation Identifiers Names and Codes (LOINC), as appropriate.

### Study Design and Participants

Cases were defined as patients with **bilateral CAS or occlusion**, identified by the ICD-10-CM code **I65.23** (Occlusion and stenosis of bilateral carotid arteries).

Controls were patients with **documented atherosclerosis but without CAS or occlusion**, identified by ICD-10-CM codes in the **I70** category (Atherosclerosis; e.g., I70.0, I70.2-, I25.10) or other relevant atherosclerosis codes, while explicitly excluding any codes in the I65.2-series (occlusion and stenosis of precerebral arteries, including carotid).

### Eligibility Criteria

Patients were excluded if they had any of the following ICD-10-CM (or equivalent procedure/pharmacy) codes recorded at any time: Transient ischemic attack (TIA): **G45.9**, Ischemic stroke: **I63.-**, Hemorrhagic stroke: **I60.-** or **I61.-**, Carotid endarterectomy (CEA): Relevant ICD-10-PCS codes (e.g., 03CH0ZZ–03CK0ZZ), Intravenous thrombolysis: Relevant ICD-10-PCS codes (e.g., 3E03317), Endovascular thrombectomy (EVT): Relevant ICD-10-PCS codes for intracranial vessels, ACA or MCA occlusion: **I66.1-** (ACA) or **I66.0-** (MCA), Iron deficiency: **D50.-** (supplemented by laboratory criteria where available: ferritin <30 ng/mL and transferrin saturation <20%), And current use of antipsychotic medications: Identified via NDC codes

### Matching Procedure

Using the TriNetX platform’s built-in propensity score matching tool, cases and controls were 1:1 matched via nearest-neighbor greedy matching with a caliper of 0.1 standard deviations of the propensity score logit. Matching covariates included: Age, Race and ethnicity, Chronic kidney disease (CKD) (**N18.-**), Neuropathy (**G62.-** or diabetes-related codes), PAD (**I73.9** or **I70.2-**), Parkinson’s disease (PD) (**G20**), And use of selective serotonin reuptake inhibitors (SSRIs; identified via NDC codes)

### Outcome Assessment

The primary outcome was **RLS**, identified by the ICD-10-CM code **G25.81** (Restless legs syndrome) recorded in any encounter.

### Statistical Analysis

Baseline characteristics were compared before and after matching. The association between bilateral CAS/occlusion and RLS was estimated using conditional logistic regression within the TriNetX platform to account for the matched design, with results reported as odds ratios and 95% confidence intervals.

## RESULTS

Before propensity score matching, patients with bilateral carotid stenosis (n = 72,890) were older than those without carotid stenosis (n = 950,967) (mean age 72.7 ± 9.46 years vs. 68.9 ± 12.4 years) and had a higher proportion of White individuals (81.3% vs. 61.0%) and non-Hispanic or Latino ethnicity (77.4% vs. 57.8%). The prevalence of several key comorbidities was also substantially higher in the bilateral carotid stenosis group, including chronic kidney disease (22.0% vs. 12.3%) and polyneuropathy (6.8% vs. 3.3%). Notably, peripheral artery atherosclerosis was present in 19.9% of patients with bilateral carotid stenosis and in 0% of the unmatched control group. Use of selective serotonin reuptake inhibitors (SSRIs) was more frequent in the stenosis group (13.7% vs. 8.0%).

After 1:1 propensity score matching (n = 58,382 per group), the two cohorts were well-balanced across all measured covariates. Mean age was nearly identical (72.9 ± 9.49 years in the bilateral carotid stenosis group vs. 72.9 ± 9.5 years in the matched control group). Standardized differences for race/ethnicity categories were minimal (White: 81.23% vs. 81.32%; Black or African American: 8.26% vs. 8.29%; Hispanic or Latino: 5.84% vs. 5.82%), as were those for sex (male: 53.84% in both groups), chronic kidney disease (21.36% vs. 21.38%), polyneuropathy (6.40% vs. 6.27%), Parkinson’s disease (0.93% vs. 0.87%), and SSRI use (13.65% vs. 13.64%). Peripheral artery atherosclerosis remained recorded only in the stenosis group in the original data but was not used as a matching covariate in the final model.

Restless legs syndrome (RLS) was present in 1,154 patients (1.977%) in the bilateral carotid stenosis/occlusion cohort compared with 1,060 patients (1.816%) in the control cohort. This corresponded to a small but statistically significant absolute risk difference of 0.161% (95% CI: 0.005% to 0.317%; z = 2.017, p = 0.0437). The risk ratio for RLS in patients with bilateral CAS/occlusion was 1.089 (95% CI: 1.002 to 1.182), and the odds ratio was 1.09 (95% CI: 1.002 to 1.186), indicating a modestly increased likelihood of RLS in the presence of severe bilateral CSD. These findings suggest a statistically significant association between bilateral CAS or occlusion and RLS with an approximately 9% higher relative risk after accounting for matched covariates.

## DISCUSSION

In this large, propensity score-matched analysis using the TriNetX Global Health Research Network, we found a modest but statistically significant association between bilateral carotid artery stenosis or occlusion and restless legs syndrome (RLS). Patients with severe bilateral carotid disease had approximately 9% higher odds of an RLS diagnosis (OR 1.09, 95% CI 1.002–1.186) compared with matched patients with atherosclerosis but without carotid stenosis or occlusion, after rigorous exclusion of prior cerebrovascular events and other known secondary causes of RLS.

The small effect size is consistent with the multifactorial etiology of RLS, in which genetic predisposition, brain iron dysregulation, and dopaminergic dysfunction predominate.^15^ Severe bilateral CAS may contribute through chronic subclinical cerebral hypoperfusion, particularly affecting dopamine-rich regions, or through associated cerebral small-vessel disease and endothelial inflammation that impair brain iron acquisition and dopaminergic signaling.^4^ These pathways provide biological plausibility for our findings, even though the absolute increase in RLS prevalence was small (1.98% vs. 1.82%). These results provide preliminary groundwork to support further studies where more granular clinical data can shed light on this relationship.

Our results align with prior observations linking vascular risk factors to RLS. Population-based studies have reported associations between cardiovascular disease, hypertension, and RLS, though many did not adjust as rigorously for confounders.^16^ By matching on key comorbidities (CKD, peripheral neuropathy, PAD, PD, and SSRI use) and excluding patients with overt stroke or TIA, our study isolates the effect of severe CAS more effectively than most previous reports. The consistency of our findings with vascular parkinsonism literature—where lower-limb restlessness is common—further supports a cerebrovascular contribution to lower-limb movement disorders.^17^ Furthermore, are study is aligned with others showing sonographic markers of carotid artery stenosis are associated with restless legs syndrome.^18,19^

Strengths of this study include the large sample size (>58,000 matched pairs), rigorous propensity score matching, and comprehensive exclusion criteria that minimize confounding from established secondary causes of RLS. The use of real-world data from the TriNetX Global Network enhances generalizability across diverse healthcare settings. For our control group, we used patients with atherosclerosis without CAS. This is due to the fact that there is no ICD-10 code to our knowledge which coded specifically for an absence of CAS. However, given the aforementioned discussion of how atherosclerosis is an independent risk factor for RLS, this may help further adjust for confounders given that we were able to include patients with AD but no CAS using a single diagnostic code.

Several limitations warrant consideration. First, reliance on ICD-10 coding (G25.81 for RLS) may introduce misclassification; this code has moderate sensitivity and specificity for clinically confirmed RLS and likely captures only more symptomatic cases. Although we excluded iron deficiency using diagnostic codes and laboratory criteria where available, residual undiagnosed brain iron deficiency could confound the association. TriNetX data lack detailed clinical phenotyping (e.g., RLS severity, polysomnography, or carotid imaging severity), limiting mechanistic insights. Finally, despite matching, unmeasured confounding (e.g., smoking, obesity, or genetic RLS risk variants) remains possible.

Our study sets the groundwork for future investigations. The question that naturally follows from our study is whether carotid revascularization procedures would have an impact on RLS. Clinically, this could be of interest in patients with medically-refractory RLS, which in one study was a substantial 17.4% of their cohort.^20^ Moreover, medical treatments for RLS, such as dopamine agonists and opioids, can be either poorly tolerated or contraindicated in certain patients.^21^ Given the relatively small absolute increase in RLS effect size, it would be interesting to investigate whether CAS produces a synergistic effect with other RLS risk factors.

## CONCLUSION

In this large propensity score-matched analysis utilizing data from the TriNetX Global Health Research Network (N=116,764), bilateral carotid artery stenosis or occlusion was associated with a modest but statistically significant increase in restless legs syndrome (RLS; odds ratio 1.09, 95% CI 1.002–1.186; p=0.0437), corresponding to an absolute risk difference of 0.161%. After excluding prior stroke, iron deficiency, antipsychotic use, and other established secondary causes, and matching for age, race/ethnicity, CKD, peripheral neuropathy, PAD, PD, and SSRI use, these findings suggest that severe bilateral carotid disease may contribute to secondary RLS through chronic subclinical hypoperfusion or microemboli affecting dopaminergic subcortical structures. To our knowledge, this represents the first demonstration of this association independent of overt stroke. Despite limitations including reliance on diagnostic coding and potential residual confounding, the results highlight a potential vascular mechanism in RLS pathogenesis, potentially shared with vascular parkinsonism, and warrant prospective studies to evaluate dose-dependency, the impact of unilateral disease, and whether carotid revascularization ameliorates symptoms in patients with refractory RLS and concurrent severe CAS.

## Data Availability

All data used in the preparation of this manuscript is available publicly and on request.

https://trinetx.com/

## Appendix ICD-10 Codes Used in Study Definitions

### 1. Case Definition (Bilateral Carotid Artery Stenosis/Occlusion)

- **I65.23**: Occlusion and stenosis of bilateral carotid arteries (This is the specific code used to identify cases with bilateral involvement.)

### 2. Control Definition (Atherosclerosis without Carotid Stenosis/Occlusion)

- **I70.-** (Atherosclerosis category, excluding I65.2-series for precerebral arteries/carotid) Examples include:
- **I70.0**: Atherosclerosis of aorta
- **I70.2-**: Atherosclerosis of native arteries of extremities (e.g., I70.20-, I70.21-, I70.22-, etc., for various sites and complications like ulceration or gangrene)
- **I70.90**: Unspecified atherosclerosis
- **I25.10**: Atherosclerotic heart disease of native coronary artery without angina pectoris (mentioned as an example of relevant atherosclerosis codes) (Controls required presence of I70 category or similar atherosclerosis codes, with explicit exclusion of any I65.2-codes.)

### 3. Exclusion Criteria

- **Transient ischemic attack (TIA)**: **G45.9** — Transient cerebral ischemic attack, unspecified (Broader category: G45.- Transient cerebral ischemic attacks and related syndromes)
- **Ischemic stroke**: **I63.-** — Cerebral infarction (Includes subcodes for thrombosis, embolism, unspecified, etc., by artery/location)
- **Hemorrhagic stroke**:
  - **I60.-** — Nontraumatic subarachnoid hemorrhage
  - **I61.-** — Nontraumatic intracerebral hemorrhage (Examples: I61.0 Nontraumatic intracerebral hemorrhage in hemisphere, subcortical)
- **Carotid endarterectomy (CEA)**: Relevant ICD-10-PCS codes (e.g., **03CH0ZZ– 03CK0ZZ** range for extirpation or repair procedures on carotid arteries; specific codes vary by approach, laterality, and device)
- **Intravenous thrombolysis**: Relevant ICD-10-PCS codes (e.g., **3E03317** — Introduction of other thrombolytic into peripheral vein, percutaneous approach; broader 3E03* for peripheral vein introductions of thrombolytics)
- **Endovascular thrombectomy (EVT)**: Relevant ICD-10-PCS codes for intracranial vessels (e.g., **03CG3ZZ** — Extirpation of matter from intracranial artery, percutaneous approach; **03CH3ZZ, 03CJ3ZZ, 03CK3ZZ** for specific arteries)
- **Anterior cerebral artery (ACA) occlusion**: **I66.1-** — Occlusion and stenosis of anterior cerebral artery
- **Middle cerebral artery (MCA) occlusion**: **I66.0-** — Occlusion and stenosis of middle cerebral artery
- **Iron deficiency**: **D50.-** — Iron deficiency anemia (Supplemented by lab criteria: ferritin <30 ng/mL and transferrin saturation <20%)
- **Current use of antipsychotic medications**: Identified via National Drug Codes (NDC); no specific ICD-10-CM code listed

### 4. Matching Covariates

- **Age**: Continuous variable (no ICD code)
- **Race and ethnicity**: Demographic variables (no ICD code)
- **Chronic kidney disease (CKD)**: **N18.-** — Chronic kidney disease (includes stages)
- **Neuropathy**: **G62.-** — Other and unspecified polyneuropathies (Or diabetes-related codes, e.g., E11.- with neurological complications)
- **Peripheral artery disease (PAD)**: **I73.9** — Peripheral vascular disease, unspecified **or I70.2-** — Atherosclerosis of native arteries of extremities
- **Parkinson’s disease (PD)**: **G20** — Parkinson’s disease
- **Selective serotonin reuptake inhibitors (SSRIs)**: Identified via National Drug Codes (NDC); no specific ICD-10-CM code

### 5. Primary Outcome

- **Restless legs syndrome (RLS)**: **G25.81** — Restless legs syndrome

**Notes**:

- All diagnoses used ICD-10-CM codes unless otherwise specified (e.g., procedures via ICD-10-PCS, medications via NDC).
- The study excluded any codes in the **I65.2-** series (occlusion and stenosis of precerebral arteries, including carotid) from controls to ensure no overlap with case definition.
- Laboratory criteria (e.g., ferritin/transferrin saturation for iron deficiency) supplemented ICD codes where noted.
- This appendix is not exhaustive of all possible subcodes but captures the ranges and examples explicitly or implicitly referenced in the Methods.

---

**Table 1:**
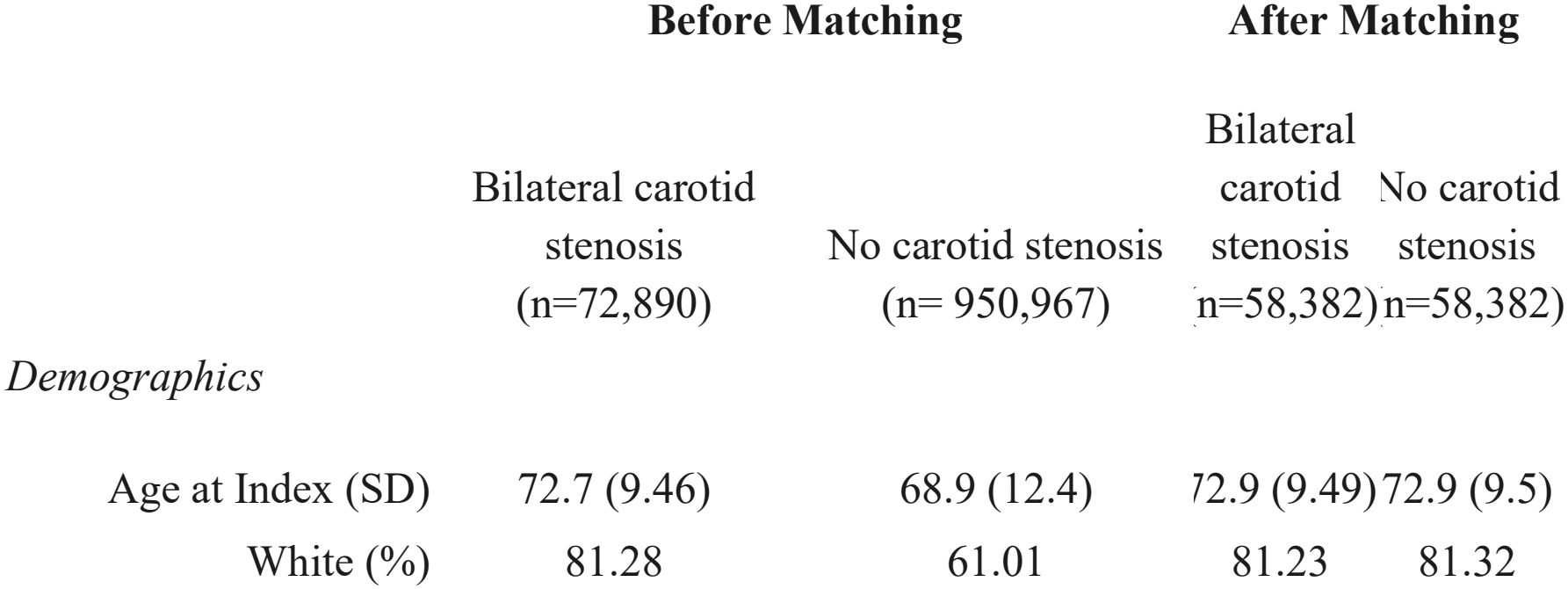
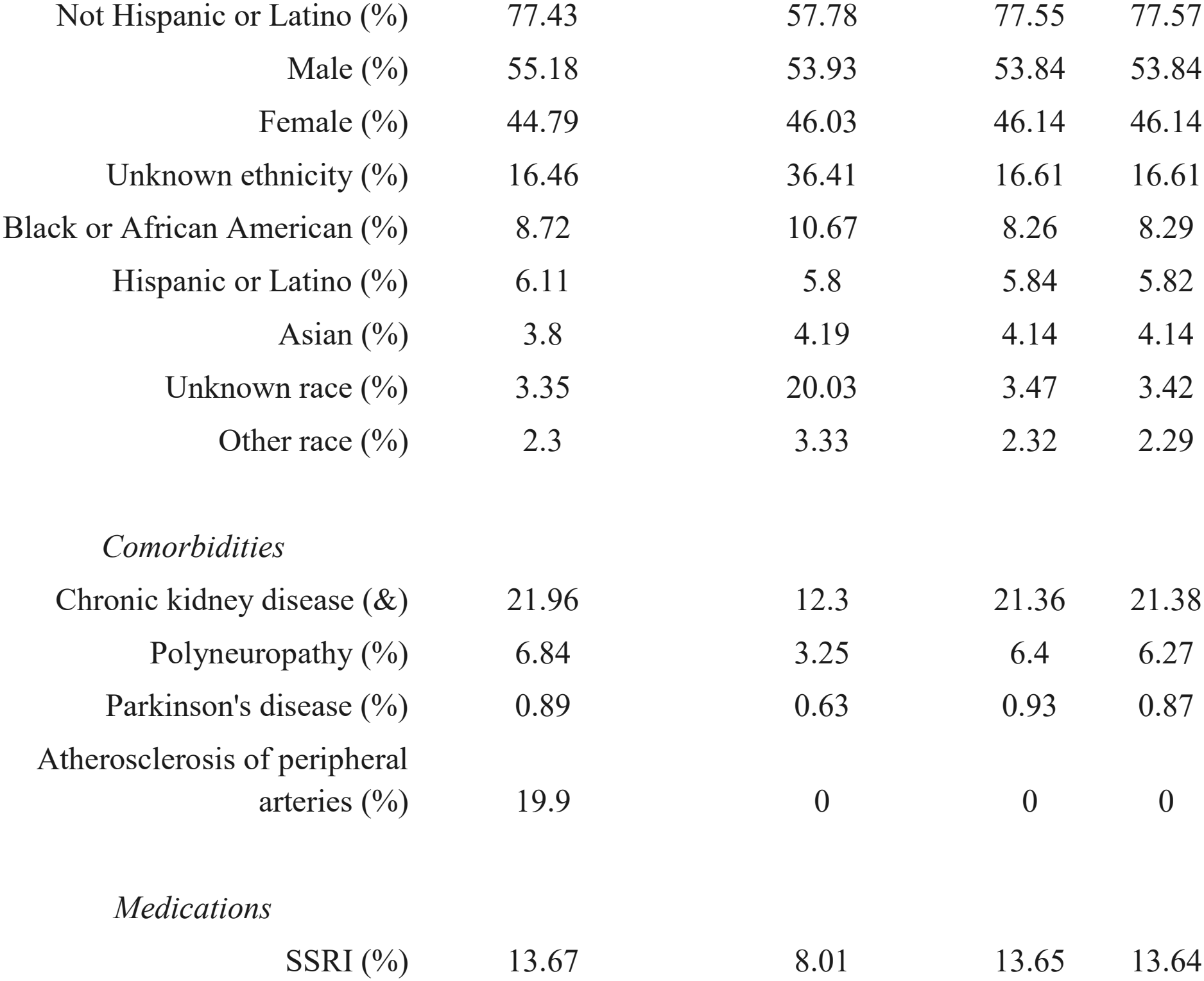
Prevalence of demographics, comorbidities and medications used as covariates for analysis before and after matching.

|  | **Before Matching** |  | **After Matching** |  |
|---|---|---|---|---|
|  | Bilateral carotid stenosis (n=72,890) | No carotid stenosis (n= 950,967) | Bilateral carotid stenosis (n=58,382) | No carotid stenosis (n=58,382) |
| *Demographics* |  |  |  |  |
| Age at Index (SD) | 72.7 (9.46) | 68.9 (12.4) | 72.9 (9.49) | 72.9 (9.5) |
| White (%) | 81.28 | 61.01 | 81.23 | 81.32 |
| Not Hispanic or Latino (%) | 77.43 | 57.78 | 77.55 | 77.57 |
| Male (%) | 55.18 | 53.93 | 53.84 | 53.84 |
| Female (%) | 44.79 | 46.03 | 46.14 | 46.14 |
| Unknown ethnicity (%) | 16.46 | 36.41 | 16.61 | 16.61 |
| Black or African American (%) | 8.72 | 10.67 | 8.26 | 8.29 |
| Hispanic or Latino (%) | 6.11 | 5.8 | 5.84 | 5.82 |
| Asian (%) | 3.8 | 4.19 | 4.14 | 4.14 |
| Unknown race (%) | 3.35 | 20.03 | 3.47 | 3.42 |
| Other race (%) | 2.3 | 3.33 | 2.32 | 2.29 |
| *Comorbidities* |  |  |  |  |
| Chronic kidney disease (&) | 21.96 | 12.3 | 21.36 | 21.38 |
| Polyneuropathy (%) | 6.84 | 3.25 | 6.4 | 6.27 |
| Parkinson's disease (%) | 0.89 | 0.63 | 0.93 | 0.87 |
| Atherosclerosis of peripheral arteries (%) | 19.9 | 0 | 0 | 0 |
| *Medications* |  |  |  |  |
| SSRI (%) | 13.67 | 8.01 | 13.65 | 13.64 |

**Table 2.**
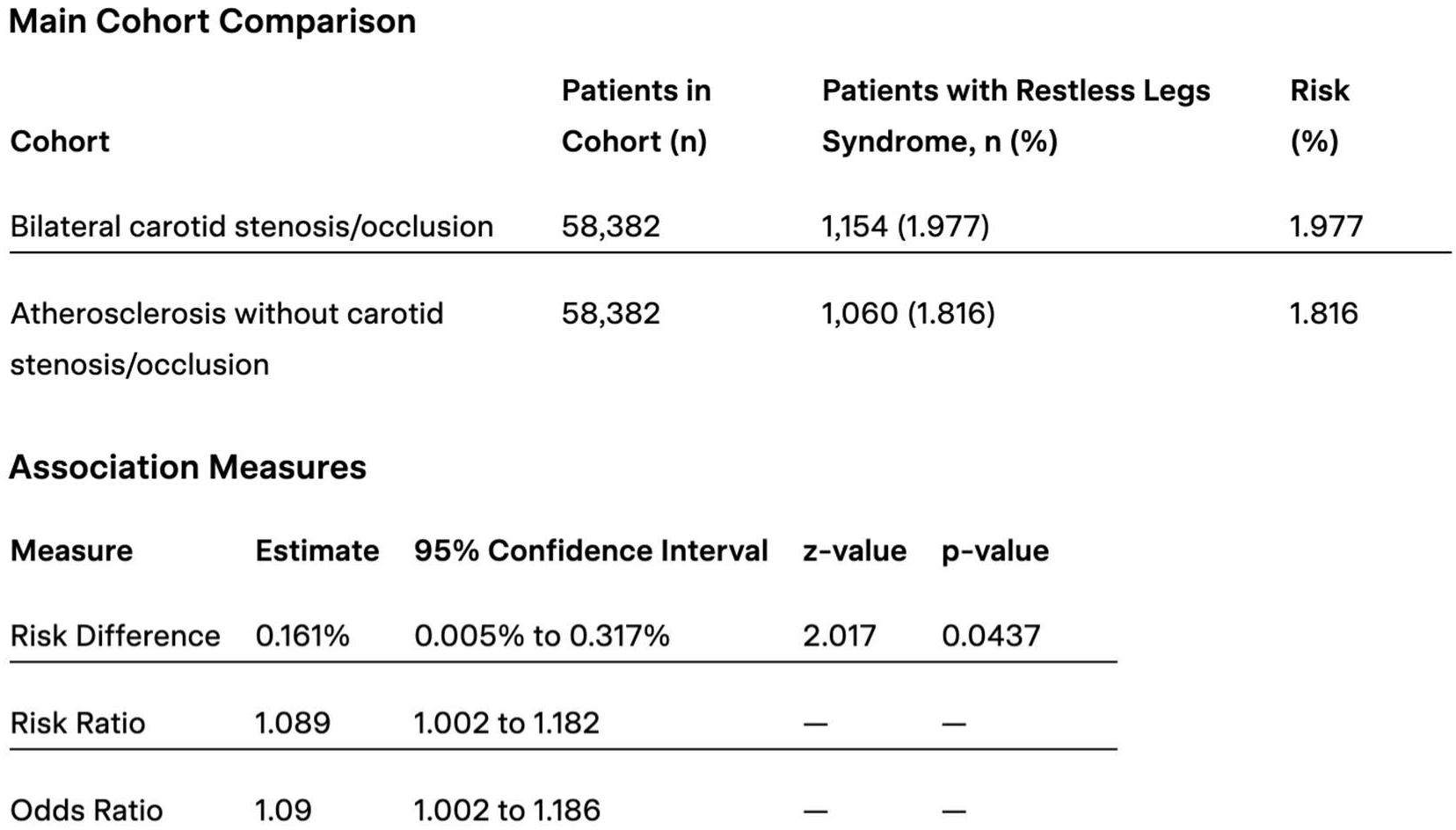
Association Between Bilateral Carotid Artery Stenosis/Occlusion and Restless Legs Syndrome in Propensity Score–Matched Cohorts.

| Main Cohort Comparison |  |  |  |  |
|---|---|---|---|---|
| Cohort | Patients in Cohort (n) | Patients with Restless Legs Syndrome, n (%) | Risk (%) |  |
| Bilateral carotid stenosis/occlusion | 58,382 | 1,154 (1.977) | 1.977 |  |
| Atherosclerosis without carotid stenosis/occlusion | 58,382 | 1,060 (1.816) | 1.816 |  |
| **Association Measures** |  |  |  |  |
| **Measure** | **Estimate** | **95% Confidence Interval** | **z-value** | **p-value** |
| Risk Difference | 0.161% | 0.005% to 0.317% | 2.017 | 0.0437 |
| Risk Ratio | 1.089 | 1.002 to 1.182 | — | — |
| Odds Ratio | 1.09 | 1.002 to 1.186 | — | — |

## REFERENCES

(1) Garcia-Borreguero D, Egatz R, Winkelmann J, Klaus B. Epidemiology of restless legs syndrome: The current status. Sleep Medicine Reviews. 2006;10(3):153–167. doi:10.1016/j.smrv.2006.01.001

(2) Park, J. H., Razuk, A., Saad, P. F., Telles, G. J. P., Karakhanian, W. K., Fioranelli, A., Rodrigues, A. C., Volpiani, G. G., Campos, P., Yamada, R. M., Castelli, V., & Caffaro, R. A. (2012a). Carotid stenosis: What is the high-risk population? Clinics, 67(8), 865–870. 10.6061/clinics/2012(08)02

(3) Walters, A. S., & Rye, D. B. (2009). Review of the relationship of restless legs syndrome and periodic limb movements in sleep to hypertension, heart disease, and stroke. Sleep, 32(5), 589–597. PubMed Central. 10.1093/sleep/32.5.589

(4) Higuchi, T., Abe, M., Mizuno, M., Yamazaki, T., Suzuki, H., Moriuchi, M., Oikawa, O., Okawa, E., Ando, H., & Okada, K. (2015). Association of restless legs syndrome with oxidative stress and inflammation in patients undergoing hemodialysis. Sleep Medicine, 16(8), 941–948. 10.1016/j.sleep.2015.03.025

(5) Gao, X., Ba, D. M., Bagai, K., Liu, G., Ma, C., & Walters, A. S. (2021). Treating restless legs syndrome was associated with low risk of cardiovascular disease: A cohort study with 3.4 years of follow-up. Journal of the American Heart Association, 10(4), e018674. PubMed Central. 10.1161/JAHA.120.018674

(6) Winter, A. C., Schürks, M., Glynn, R. J., Buring, J. E., Michael, G. J., Klaus, B., & Kurth, T. (2013). Vascular risk factors, cardiovascular disease, and restless legs syndrome in women. The American Journal of Medicine, 126(3), 220–227.e2. 10.1016/j.amjmed.2012.06.040

(7) Sharkey-Toppen, T. P., Tewari, A. K., & Raman, S. V. (2014). Iron and atherosclerosis: Nailing down a novel target with magnetic resonance. Journal of Cardiovascular Translational Research, 7(5), 533–542. PubMed Central. 10.1007/s12265-014-9551-y

(8) Allen, R. P., Auerbach, S., Bahrain, H., Auerbach, M., & Earley, C. J. (2013). The prevalence and impact of restless legs syndrome on patients with iron deficiency anemia. American Journal of Hematology, 88(4), 261–264. 10.1002/ajh.23397

(9) Makwana, A., Sablok, R., Adler, P., Appireddy, M. R., Philip, A., & Shukla, G. (2026). Phenotypes and course of restless legs syndrome in acute stroke survivors – a prospective observational cohort study. Sleep Medicine, 138, 108709. 10.1016/j.sleep.2025.108709

(10) Rice, M. E., Patel, J. C., & Cragg, S. J. (2011). Dopamine release in the basal ganglia. Neuroscience, 198, 112–137. PubMed Central. 10.1016/j.neuroscience.2011.08.066

(11) Kuzucu, P., Çeltikçi, P., Demirtaş, O. K., Canbolat, Ç., Çeltikçi, E., Demirci, H., Özışık, P., Tubbs, R. S., Pamir, M. N., & Güngör, A. (2023). Arterial supply of the basal ganglia: A fiber dissection study. Operative Neurosurgery, 24(5), e351–e359. 10.1227/ons.0000000000000612

(12) Finn, C., Giambrone, A. E., Gialdini, G., Delgado, D., Baradaran, H., Kamel, H., & Gupta, A. (2017). The association between carotid artery atherosclerosis and silent brain infarction: A systematic review and meta-analysis. Journal of Stroke and Cerebrovascular Diseases : The Official Journal of National Stroke Association, 26(7), 1594–1601. 10.1016/j.jstrokecerebrovasdis.2017.02.028

(13) Gu, T., Hedberg, I., Vikner, L., Björnebäck, A., Karlsson, J., Henze, A., Fox, A. J., & Johansson, E. (2025). Incidence of microembolic signals on transcranial doppler among patients with symptomatic carotid near occlusion. European Journal of Vascular and Endovascular Surgery, 69(4), 522–530. 10.1016/j.ejvs.2024.11.012

(14) Nassar, M., Abosheaishaa, H., Elfert, K., Beran, A., Ismail, A., Mohamed, M., Misra, A., Essibayi, M. A., Altschul, D. J., & Azzam, A. Y. (2025). TriNetX and real-world evidence: A critical review of its strengths, limitations, and bias considerations in clinical research. ASIDE Internal Medicine, 1(2), 24–33. PubMed Central. 10.71079/aside.im.03222516

(15) Allen, R. P. (2007). Controversies and challenges in defining the etiology and pathophysiology of restless legs syndrome. Recognizing and Treating Restless Legs Syndrome: Current Standards, 120(1, Supplement 1), S13–S21. 10.1016/j.amjmed.2006.11.003

(16) Giannini, G., Zanigni, S., Melotti, R., Gögele, M., Provini, F., Facheris, M. F., Cortelli, P., & Pramstaller, P. P. (2014). Association between restless legs syndrome and hypertension: A preliminary population-based study in South Tyrol, Italy. European Journal of Neurology, 21(1), 72–78. PubMed. 10.1111/ene.12244

(17) Korczyn, A. D. (2015). Vascular parkinsonism—Characteristics, pathogenesis and treatment. Nature Reviews Neurology, 11(6), 319–326. 10.1038/nrneurol.2015.61

(18) Say, A., & İşcan, D. (2025). An evaluation of carotid doppler indices in patients with restless legs syndrome. Medicine, 104(30), e43302. PubMed Central. 10.1097/MD.0000000000043302

(19) Zolfaghari, S., Dasgupta, K., & Postuma, R. B. (2020). Restless leg syndrome and objectively-measured atherosclerosis in the Canadian longitudinal study on aging. Movement Disorders, 35(12), 2314–2318. 10.1002/mds.28326

(20) Petramfar, P., & Jankovic, J. (2024). Medication refractory restless legs syndrome: Real-world experience. Journal of the Neurological Sciences, 463, 123121. PubMed. 10.1016/j.jns.2024.123121

(21) Wei, S., Song, X., Chen, R., Chen, S., & Lu, B. (2025). Exploring the top 50 drugs associated with restless legs syndrome based on the FDA data from 2004 to 2024. Nature and Science of Sleep, 17, 929–946. PubMed Central. 10.2147/NSS.S513538

